# Enhanced Direct Joint Attenuation and Scatter Correction of Whole-Body PET Images via Context-Aware Deep Networks

**DOI:** 10.1101/2022.05.26.22275662

**Authors:** Saeed Izadi, Isaac Shiri, Carlos F. Uribe, Parham Geramifar, Habib Zaidi, Arman Rahmim, Ghassan Hamarneh

## Abstract

In positron emission tomography (PET), attenuation and scatter corrections is necessary steps towards accurate quantitative reconstruction of the radiopharmaceutical distribution. Inspired by recent advances in deep learning, many algorithms based on convolutional neural networks have been proposed for automatic attenuation and scatter correction, enabling applications to CT-less or MR-less PET scanners to improve performance in the presence of CT-related artifacts. A known characteristic of PET imaging is to have varying tracer uptakes for various patients and/or anatomical regions. However, existing deep learning-based algorithms utilize a fixed model across different subjects and/or anatomical regions during inference, which could result in spurious outputs. In this work, we present a novel deep learning-based framework for direct reconstruction of attenuation and scatter corrected PET from non-attenuation-corrected images in absence of structural information. To deal with inter-subject and intra-subject uptake variations in PET imaging, we propose a novel model to perform subject- and region-specific filtering through modulating the convolution kernels in accordance to the contextual coherency within the neighboring slices. This way, the context-aware convolution can guide the composition of intermediate features in favor of regressing input-conditioned and/or region-specific tracer uptakes. We also utilize a large cohort of 910 whole-body studies for training and evaluation purposes, which is more than one order of magnitude larger than previous works. In our experimental studies, qualitative assessments showed that our proposed CT-free method is capable of producing corrected PET images that accurately resemble ground truth images corrected with the aid of CT scans. For quantitative assessments, we evaluated our proposed method over 112 held-out subjects and achieved absolute relative error of 14.30 ± 3.88% and relative error of − 2.11% ± 2.73% in whole-body.

## 1. Introduction & Related Work

^18^F-fluorodeoxyglucose (FDG) positron emission tomography (PET) imaging is one of the leading imaging modalities for quantitative *in vivo* measurement of physiological and biochemical processes with applications in oncology (Rohren et al., 2004), cardiology (Dilsizian et al., 2016) and neurology (Herholz and Heiss, 2004). Studies reveal that 40-50% of the recorded annihilation coincidences in a typical whole-body PET scan are affected by Compton scatter where one or both photons deviate from the original line of response before reaching the PET detectors (Zaidi and Koral, 2004). The scattered photons may interact with other surrounding dense material, such as bed instruments, electronic components, and patient body, and carry inaccurate information about the annihilation site (Gong et al., 2019). Also, scattered photons may fall out of the predefined energy windows and fail to be recorded by the PET detectors, which leads to photon attenuation (Gong et al., 2019). In either case, inaccurate number of detected coincidences contributes to under-estimation or over-estimation of the tracer distribution, which ultimately results in inaccurate uptake quantification and notable visual artifacts in the reconstructed PET images. These factors can complicate the assessment of PET findings and may lead to incorrect (false negative/positive) clinical diagnoses. Accordingly, it is crucial to correct for the loss of annihilation photons (attenuation correction) and the inaccurately recorded coincidences (scatter correction) during image reconstruction (Boellaard, 2009).

With the introduction of PET/CT (Kinahan et al., 1998), and more recently PET/MR scanners (Fei et al., 2009), high-resolution anatomical images from MRI and CT images are leveraged to derive attenuation and scatter information to obtain attenuation-and-scatter-corrected PET images (PET-AC) (Gong et al., 2019). In hybrid PET/CT scanners, the Hounsfield units recorded in CT scans can be directly converted into the PET 511-keV linear attenuation coefficients. Despite their popularity in routine clinical practice, the CT-based correction methods have several drawbacks including the risk of artifact propagation and position mismatch between PET and CT scans (Berker and Li, 2016) and the exposure to ionizing radiation; of particular concern for pregnant and pediatric patients (Cheuk et al., 2012).

Alternatively, PET/MR is a non-ionizing imaging technique that allows enhanced visualization of soft tissue contrast without necessitating radiation exposure (Vandenberghe and Marsden, 2015). However, aside from the relatively more limited availability of MR scanning time, the MRI tissue intensities cannot be directly converted into 511 keV linear attenuation coefficients as in CT-based systems (Yang et al., 2019b). To leverage MRI for PET attenuation correction, segmentation and registration-based techniques have been developed. In segmentation-based techniques, uniform linear attenuation coefficients are assigned to different tissues inferred from segmenting an MRI image (Arabi et al., 2015; Berker et al., 2012). In the registration-based methods, an atlas of pre-acquired CT images is used to obtain an attenuation map template, which is then spatially registered to match the patient’s body using the anatomical information in the MRI (Arabi et al., 2016). In addition to the computational overhead, these techniques may deteriorate PET reconstruction due to tissue mis-classification, imprecise co-registration, data truncation and metal-induced susceptibility artifacts in CT and/or MR (Lillington et al., 2020; Gong et al., 2019).

Furthermore, enabling CT/MRI-less PET scanners can open up new possibilities for more compact, affordable PET-only scanners of the future (e.g. used for screening, etc.). To avoid relying on structural information captured by CT or MRI, an attempts was made to simultaneously reconstruct the tracer distribution and attenuation maps from the emission data only using maximum likelihood estimation of uptake and attenuation (MLAA) (Nuyts et al., 1999). However, MLAA suffers from slow convergence and low signal-to-noise ratio (SNR) (Mehranian and Zaidi, 2015). Advances in electronics and scintillation research have augmented some commercial PET scanners with time-of-flight (TOF) information. The use of TOF information expedites the convergence time of MLAA and improves SNR (Defrise et al., 2012). Nevertheless, the shortcomings of existing TOF-based algorithms persist mainly due to the uncertainty in detecting the true event positions given the limited time resolution of TOF and high noise.

In recent years, the popularity of deep learning (DL) has ignited extensive research aimed at leveraging the capabilities of neural networks in medical imaging (Litjens et al., 2017; Yedder et al., 2021; Zaidi and El Naqa, 2021). The representative works for attenuation and scatter correction (ASC) using convolution neural networks (CNN) include generation of pseudo-CT images from MR sequences such as ultra-short echo time (UTE) (Leynes et al., 2018), zero echo time (ZTE) (Gong et al., 2018; Blanc-Durand et al., 2019), and Dixon sequences (Leynes et al., 2018). There have been some recent efforts to directly produce pseudo-CT images from non-attenuation-corrected images (PET-NC) (Dong et al., 2019) or uptake and attenuation maps obtained from MLAA (Hwang et al., 2018, 2019). In addition, several studies have employed more advanced DL-based techniques, such as generative adversarial networks (GAN), to boost the performance of pseudo-CT synthesis (Choi and Lee, 2018; Tao et al., 2020). Specifically, MedGAN framework (Armanious et al., 2020b) was utilized to approximate PET-NC to pseudo-CT mapping for whole-body images using paired training data (Armanious et al., 2020a). Recently, methods based on 3D cycleGAN have been applied to pseudo-CT synthesis with the goal of circumventing the need for paired training sets and improving pseudo-CT images by enforcing inverse consistency (Lei et al., 2020a). Despite their substantial improvements over traditional algorithms, methods based on GAN are prone to hallucinating imprecise features in the images (Cohen et al., 2018) and consequently deteriorate the performance of the downstream ASC task (Seith et al., 2017).

Another family of approaches that have recently thrived in tackling direct ASC is emission-only (PET-NC to PET-AC) techniques. These approaches only operate on the PET images or sinograms without requiring any anatomical information. The merits of the encoder-decoder networks in direct PET-NC to PET-AC mapping was demonstrated for brain (Shiri et al., 2019) and whole-body (Shiri et al., 2020) images. Most recently, Arabi et al. (Arabi and Zaidi, 2020) trained a multi-input network to predict the attenuation coefficient factors for a reference slice from its different TOF sinogram bins pertinent to the same slice. Built upon the cycleGAN framework, some works proposed to improve PET-NC to PET-AC mapping by restricting the search space through introducing a reverse network for PET-AC to PET-NC (Dong et al., 2020; Lei et al., 2020b).

Most of the DL-based algorithms for direct attenuation correction adopt either 2D or 3D networks. In the former, either individual 2D patches or full-resolution slices along the axial dimension are processed by a sequence of 2D convolutional layers while in the latter the entire volumetric input is manipulated via 3D convolutional layers during the training and inference. Even though 2D models generally benefit from relatively larger training data (i.e. slices), they fail to integrate the contextual coherency within neighboring slices. On the other hand, 3D networks provide the added value of accessing cross-slice contextual information at the expense of fewer training samples, higher computational cost, and increased number of learnable parameters that can amplify the chances of over-fitting. Furthermore, the information from distant organs are often needlessly aggregated when volumetric inputs are processed by 3D networks. An attempt has been made to use 2.5D training schemes for low-dose PET denoising where multiple consecutive slices are fed into the network to account for cross-slice contextual information (Xu et al., 2017). However, a potential drawback of 2.5D networks is that the contextual information is aggregated in the very first layer of the network and thus it is less likely for such information to influence deeper layers that have more direct impact on the final reconstruction outcome.

Another prominent characteristic of PET images is that the distribution of the tracer uptake tends to undergo considerable inter-subject variations due to changes in factors such as the administered dose, imaging time, and reconstruction parameters. Even the imaging noise and partial volume effects can also magnify the tracer variations across subjects (Gong et al., 2019). On the other hand, the tracer uptake may also vary substantially from one organ to another, even though it is nearly homogeneous inside individual anatomical regions. Despite such input-dependent variations, the existing DL-based methods adopt traditional convolution layers to solve ASC problem wherein the learned convolution kernels are kept fixed over different subjects and body organs during training and inference.

Recent research in dynamic neural networks (Jia et al., 2016; Su et al., 2019; Lin et al., 2020) and guided filtering (Tang et al., 2020; Wang et al., 2018) are promising attempts to produce context-aware networks. To perform object/activity recognition from natural images and machine translation, Lin et al. (Lin et al., 2020) proposed context-gated convolutions to adapt the convolution kernels based on the statistics of the incoming feature representation. Inspired by these works, we provide an alternative way to incorporate adequate 3D contextual information into 2D networks for CT/MRI-less ASC through introducing context-aware convolutional layers (CAC). Our proposed model differs from what mentioned above by (1) the 3D contextual information within neighboring slices is used to adaptively modulate the kernels in specific layers and the network is still in 2D manner and therefore enjoys accessing to large training instances as well as light computation cost at inference. In particular, the neighboring slices are fed into a shared sub-network to extract the region-wise information, which is subsequently used to modify the convolution kernels in an adaptive manner. We use context-aware convolutions only in the decoder layers instead of the entire network to achieve superior performance and efficiency trade-off. Patch-based training does not respect the physics of the attenuation and scatter artifacts and therefore our proposed network operates on full-resolution 2D slices in the input layer.

We address the limitations of previous PET-only ACS works by making the following contributions.

- In contrast to 3D and 2.5D networks, we propose a 2D network that exploits guidance from neighboring slices to propagate input-conditioned context in the convolution kernels.
- We propose to adopt a shared sub-network to extract context information from the set of neighboring slices, which is then used for convolution modification.
- We leverage channel and spatial interactions to adapt the convolution kernels based on the extracted contextual information.
- We conduct our study on a large cohort of 910 subjects that is more than one order of magnitude larger than previous works (Shi et al., 2019; Yang et al., 2019a; Hwang et al., 2019; Lei et al., 2020b).

## 2. Materials

### 2.1. Subjects and PET/CT Acquisition

Whole-body ^18^F-FDG PET/CT scan data of 910 subjects (demographics shown in Table 1) were acquired between 2016-2018 on a Siemens Biograph 6 True point scanner. To avoid any interventional effect on diagnosis, treatment or management of patients, retrospective use of the scan data and waiver of consent was approved by Institutional Review Board (ethic number IR.TUMS.MEDICINE.REC.1398.525) of our institute (Tehran University of Medical Science). All patients were intravenously administered 367.74 ± 48.87 of ^18^F-FDG and the PET/CT scanning was performed after an uptake period of 60 ± 13 minutes. To obtain attenuation-corrected ground truth PET images that leverage CT-based attenuation maps (PET-CT), a low-dose CT scan (110 kVp, 145 mAs) was conducted before the PET scanning. PET images were reconstructed using the ordinary Poisson ordered subsets-expectation maximization (OP-OSEM) algorithm with 2 iterations and 21 subsets followed by a 5-mm FWHM Gaussian post-reconstruction smoothing. Point-spread functions (PSF) were incorporated in the reconstruction procedure. The matrix size of the reconstructed images was 168 × 168 with a voxel size of 4.073 × 4.073 × 3 mm^2^.

**Table 1.** Patient demographics of clinical whole-body PET/CT studies enrolled in this study and train/test split information.

|  | Sex | No. | Age | Weight (kg) | Injected Dose (MBq/g) | Time Post-Injection |
| --- | --- | --- | --- | --- | --- | --- |
| | | | mean ± std<br>(min, max) | mean ± std<br>(min, max) | mean ± std<br>(min, max) | mean ± std<br>(min, max) |
| Train | Male | 422 | 48 ± 18<br>(6, 87) | 76.1 ± 16.4<br>(21.0, 145.0) | 374.7 ± 55.1<br>(123.2, 488.4) | 60 ± 14<br>(0, 238) |
| | Female | 376 | 49 ± 15<br>(2, 85) | 69.5 ± 15.0<br>(11.5, 138.0) | 369.9 ± 45.3<br>(177.6, 555.0) | 59 ± 10<br>(0, 86) |
| Test | Male | 61 | 45 ± 17<br>(11, 74) | 75.9 ± 14.4<br>(25.0, 113.0) | 366.4 ± 45.2<br>(170.2, 469.9) | 60 ± 11<br>(8, 120) |
| | Female | 51 | 50 ± 13<br>(23, 74) | 71.7 ± 12.0<br>(50.0, 106.0) | 367.0 ± 34.7<br>(284.9, 444.0) | 62 ± 17<br>(2, 160) |

### 2.2. Data Preparation

From the entire cohort, 798 subjects were randomly selected and split into training and validation cohorts with 95:5 ratio, respectively. The validation set was used to select appropriate values for training hyper-parameters and to monitor the risk of over-fitting during the training. For loss calculation, the dynamic range of the PET-NC and PET-CT images were converted to standardized uptake value (SUV) based on the injected dose, decay factor and patient’s weight available from the DICOM headers. In our implementation, we used axial slices and cropped each to 152 × 152 pixels to shrink the background (without cropping any foreground) and to emphasize the foreground tracer uptakes. For clinical evaluations, a held-out test cohort consisting of 112 subjects were used. Table 1 summarizes the patient demographics for training and test cohorts.

## 3. Methods

We first propose the mathematical formulation of direct ASC as a supervised image translation problem and introduce the necessary notations. For the solution, i.e. obtaining PET-AC outputs from PET-NC inputs, we adopt a U-Net (Ronneberger et al., 2015) architecture and equip it with the context-aware convolutional layers (CAC). The network is designed to deliver a solution with specific properties such as efficient use of contextual coherency within neighboring slices to the input 2D slice and handling inter-subject and intra-subject uptake variability.

### 3.1. Training Overview

Let 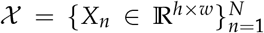 denote a whole-body 3D PET-NC image consisting of *N* axial slices each of size *h* × *w* degraded by the attenuation function Φ, and 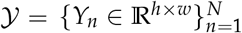 be the corresponding reference PET-CT ground truth (section 2). The attenuation degradation function for the *n*-th slice can be written as:

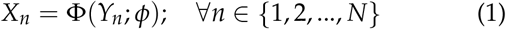

where *ϕ* indicates the set of parameters associated with the degradation function. A 2D network for direct attenuation correction aims to determine a predictive function Θ, parameterized by *θ*, which maps an arbitrary 2D slice in the PET-NC domain, i.e. *X*_*i*_, to its corresponding slice in PET-AC domain, *Y*_*i*_. In the conventional 2.5D training scheme, the network takes in a set of *M* PET-NC slices 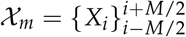 centered at *i*-th slice, known as the target slice, and produces the PET-AC counterpart. Following the formalism of a supervised training scheme and empirical risk minimization, we seek *θ*^∗^ that minimizes the distance between 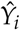 and *ϒ*_*i*_ over a training dataset.

### 3.2. Training Details

We trained our network for 5 × 10^4^ iterations with a batch size of 64. To form the training mini-batches, we firstly selected random subjects and then picked random individual 2D slice from each subject. The selected 2D slices in the mini-batch were subsequently coupled with their immediate previous and next neighbors along the axial dimension. Adam optimizer with default parameters; *β*_1_ = 0.9 and *β*_2_ = 0.999 was used for optimization. The learning rate was initially set to 7 × 10^−4^ and multiplied by 0.1 at iterations 2 × 10^4^ and 3 × 10^4^. The network parameters were initialized with the Kaiming method (He et al., 2015). We set *M* = 5 and optimize the network parameters using mean squared error (MSE).

### 3.3. Architecture Overview

Fig. 1 depicts the overview of our proposed CA-DAC network for an efficient attenuation correction. CA-DAC follows the U-Net architecture with 2*b* building blocks. In particular, we employ *b* blocks in the encoder followed by *b* symmetric blocks to form the decoder part. Every encoder block consists of two standard convolutional layers with kernel size 3 × 3 and ReLU activations. Furthermore, 2 × 2 max-pooling with stride 2 is used after each encoder block to downs-sample the feature maps until a bottleneck layer. For the decoder, transposed convolutional layer is utilized between consecutive blocks to bring the feature maps into the original input resolution. Skip connections link the encoder block to their corresponding decoder block to ensure improved information flow. We replace the standard convolutional layers in the decoder with CAC layers to incorporate the contextual information while preserving the computational efficiency. Each CAC layer takes the 2D input along with its immediate anterior and posterior neighbors, extracts information via a shared sub-network, and modifies the original convolution kernel by attention map computed through a global projection, channel interaction, and spatial interaction. Empirically, we set parameter *b* = 5 and the feature numbers in the decoder to 32, 64, 128, 256 and 512.

**Fig. 1.**
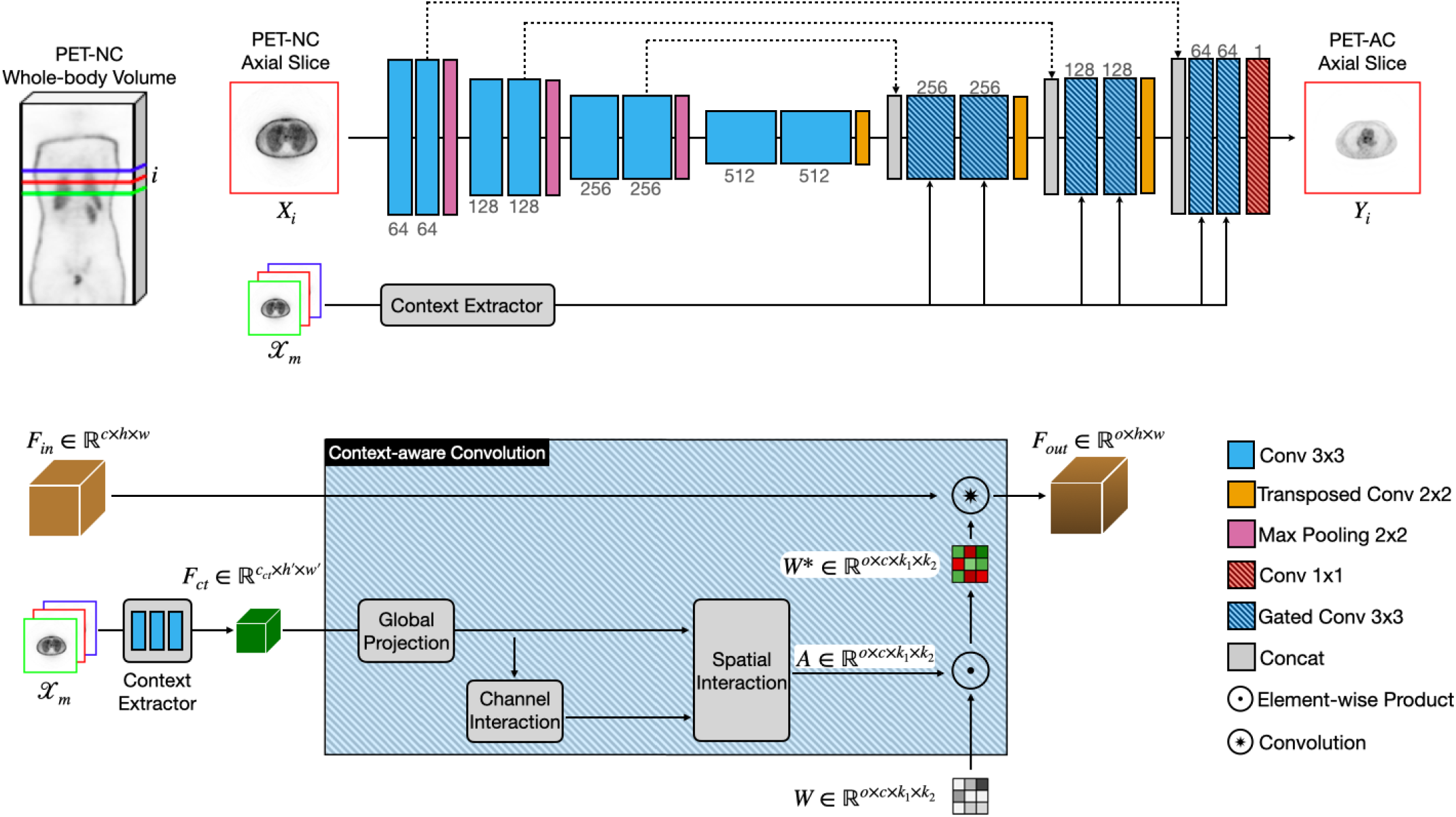
The overall architecture of our proposed network

### 3.4. Context-aware Convolution

In general, the input to a standard convolution is a feature map *F* ∈ ℝ^*c*×*h*×*w*^ with *c* input channels, height *h*, and width *w*. Then, a sliding window is swiped over the feature map to extract local patches of size 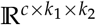 with *c* channels, height *h* and width *w*. Next, the patches are convolved with the kernel weights 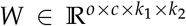 to produce the output feature maps with *o* output channels. As the weights *W* in standard convolution only depend on the local information and are agnostic to the image context, we propose CAC layers to mitigate the aforementioned issue with standard convolutions. A CAC layer learns a mapping function that outputs a context-aware attention map based on the contextual prior within neighboring input slices. The learned attention map then adaptively influences the convolution kernel in a network designed for direct ASC. Fig 2 provides a detailed illustration of CA layer.

**Fig. 2.**
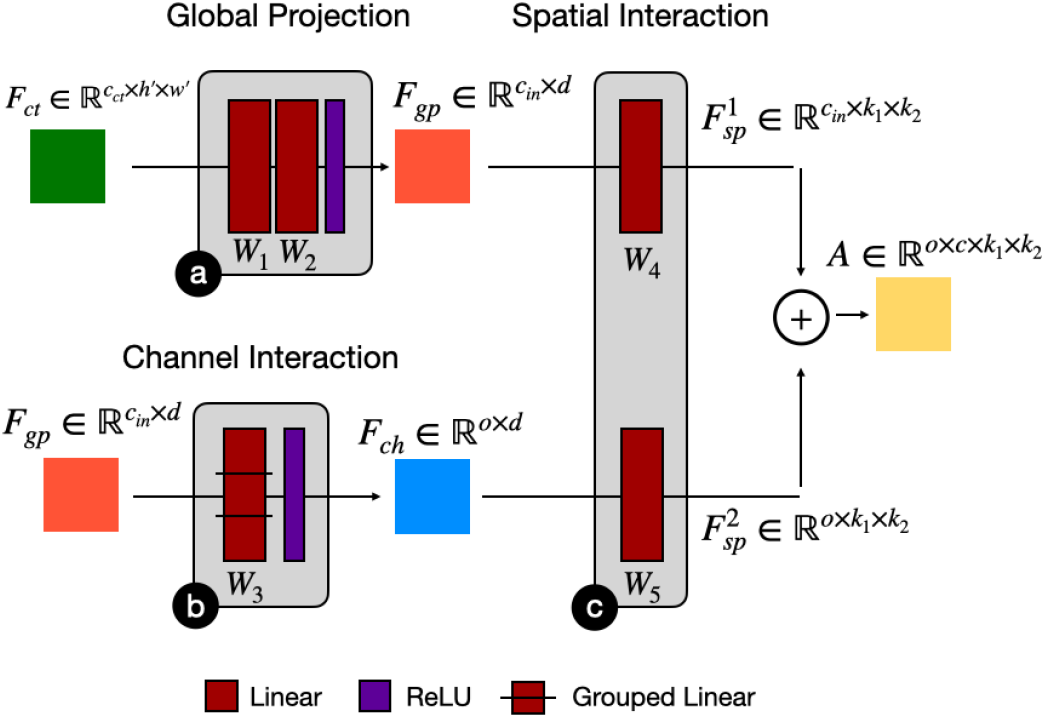
Detailed illustration of the modules in the context-aware convolutional layer.

#### Context Extractor

Here, we focus on the context extraction module. Although several choices can be considered for summarizing the contextual information in neighboring slices, we adopt a shallow network consisting of three 2-strided standard convolutional layers followed by ReLU activation for this purpose (Fig. 1-1). This network takes in the input slices 𝒳_*m*_ and extract contextual features 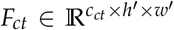 where *h*^′^ = *h*/8 and *w*^′^ = *w*/8. The extracted feature tensor *F*_*ct*_ are then leveraged in different instances of the CAC layers to modify kernel weights based on the contextual information. To improve the efficiency of the network, we keep the depth of the context extractor module to be shallow and share its weights across all instances of the CAC layers.

#### Global Projection

As the first component in a CAC layer (Fig 2-a), we use a linear layer with weights 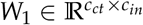 to match the number of channels in *F*_*ct*_ to the channels in the incoming features from the previous layer and obtain 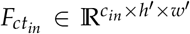. We then use another linear layer with weights *W*_2_ ∈ ℝ^*h*′*w*′×*d*^ followed by ReLU to project individual flattened channels of size *h*^′^*w*^′^ in 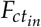 into a global latent representation 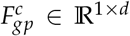. Following the bottleneck design in (Vaswani et al., 2017; Lin et al., 2020; Hu et al., 2018), we set 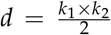. The weights *W*_2_ are shared across all channels within a CAC layer to reduce the number of learnable parameters. The output for 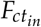 with *c*_*in*_ inputs channels is 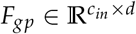.

#### Channel Interaction

The second module aims to transform *F*_*gp*_ with *d* channels to the space spanned by the desired number of output channels, i.e. *o* (Fig 2-b). Following (Lin et al., 2020), we use a grouped linear layer parametrized by 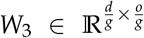 where *g* is the number of groups. The module ends with a ReLU and gives *F*_*ch*_ ∈ ℝ^*o*×*d*^ as the output.

#### Spatial Interaction

The input to the third module is both *F*_*gp*_ and *F*_*ch*_, which are decoded to the spatial size of the convolution kernel (Fig 2-c). Similar to (Lin et al., 2020), we use two linear layers with weights 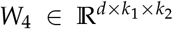 and 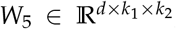, which are shared across channels in *F*_*gp*_ and *F*_*ch*_, respectively. The outputs of the module are 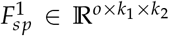 and 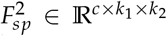. Then, the final attention map is formed by:

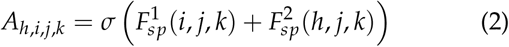

where *σ* represents the sigmoid function. Given that the original kernel 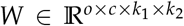 and obtained attention mask 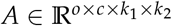 are of the same size, the underlying contextual information within the input slices can be incorporated into the updated kernel *W*^∗^ through element-wise multiplication, i.e.:

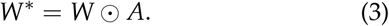

The output feature representations is then obtained by convolving the input feature maps *F* with the updated kernel *W*^∗^.

### 3.5. Evaluation Strategy

We measured the quantitative error between the reference PET-CT and predicted PET-AC outputs using relative error (RE%) and absolute relative error (ARE%) which are computed as follows:

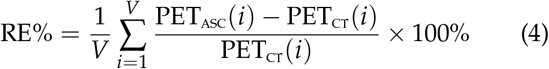

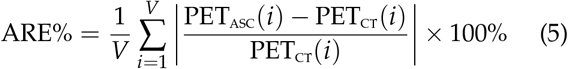

where *V* refers to the total number of voxels, and PET_ASC_ and PET_CT_ indicate the estimated PET-AC images using DL-based approaches and the CT-based ground truth, respectively. In addition to the quantification errors, popular image quality metrics including peak-signal-to-noise-ration (PSNR) and structural similarity (SSIM) were utilized to compare the visual similarity of the estimated PET-AC images against their corresponding PET-CT ground truth images. These metrics are defined as follows:

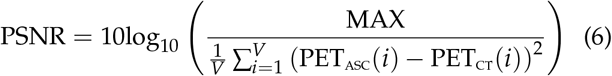

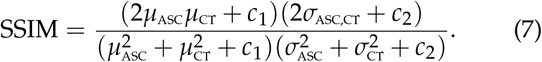

In Eq. 7, MAX refers to the maximum possible intensity value in the PET-CT image, and *µ*_ASC_ and *µ*_CT_ are the mean intensity of the PET-AC obtained by different DL-based ASC approaches and the PET-CT ground truth, respectively. Also, *σ*_ASC_ and *σ*_CT_ indicate the standard deviation in the predicted PET and CT-based ground truth images, respectively, and *σ*_DL,CT_ represents the co-variance between them. Lastly, *c*_1_ and *c*_2_ are constant values to prevent division by very small values (set to default values of *c*_1_ = 0.01 and *c*_2_ = 0.03 in our experiments).

For statistical analysis, a joint histogram was plotted to show the distribution of the measured uptake correlation between the DL-based predicted PET-AC outputs and the reference PET-CT ground truth averaged over all patients in the test cohort. We carried out all the quantitative assessments as well as the joint histogram within the SUV units (g/mL) in the range of 0.1–20.0. The SUV was calculated as follows: Image-derived uptake [MBq/mL] / injection dose [MBq] × patient’s weight [g].

## 4. Results

### 4.1. Quantitative Assessment

The performance evaluation of our proposed context-aware network (CA-DAC) includes validation against the reference PET-CT ground truth and comparison to a conventional 2.5D UNet (UN-DAC) over a held-out test cohort consisting of 112 subjects across whole-body and 6 anatomical regions. Fig 3(left) depicts the region-wise analysis of PET-AC images in terms of average RE for CA-DAC and UN-DAC. Fig 3(right) further illustrates the region-wise assessment of ARE for CA-DAC and UN-DAC approaches. Our evaluations showed that incorporating the contextual information in the decoder layers results in considerable performance gain across different anatomical regions of the body. In the whole-body, the average RE were 2.43 ± 2.94 and − 2.11 ± 2.73 for UN-DAC and CA-DAC, respectively. Also, the average ARE reduced from 14.79 ± 2.37 to 13.96 ± 2.32 for UN-DAC and CA-DAC, respectively. We noted that the highest quantification errors appeared in the chest and lung & liver regions with mean relative error of 3.11 ± 5.11 and 2.68 ± 6.40, respectively.

**Fig. 3.**
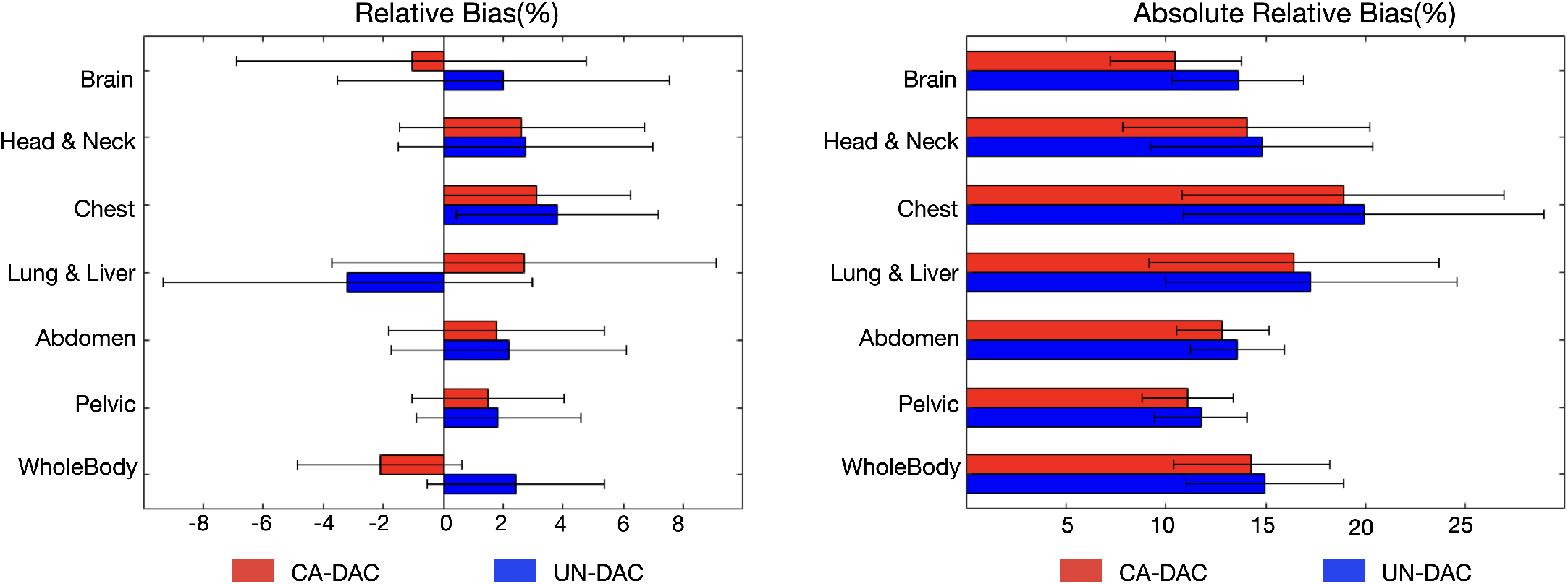
Quantitative error measured in 6 anatomical regions and whole-body for CA-DAC and UN-DAC approaches over 112 heldout subjects in terms of left) RE(%) and right) ARE(%).

This can be justified by the fact that these regions are often affected by respiratory motion artifacts during the image acquisition and thereby both their appearance and tracer uptake measurements may vary significantly across neighboring slices and ultimately lead to less coherency. The lowest error occurred in the brain region with relative error of −1.05 ± 5.83.

To quantitatively assess the visual quality of the predicted PET-AC images against the reference PET-CT images, Fig. 4 depicts the mean PSNR and mean SSIM in the different regions after attenuation correction by CA-DAC and UN-DAC. From Fig. 3, it is discernible that PET-AC outputs from CA-DAC are visually more similar to the PET-CT ground truth. In particular, we observed that CA-DAC outperformed UN-DAC by 0.71 (dB) averaged over all regions. Likewise, the SSIM scores averaged over all regions were improved from 0.9406 to 0.9465 using CA-DAC over UN-DAC in our evaluation set.

**Fig. 4.**
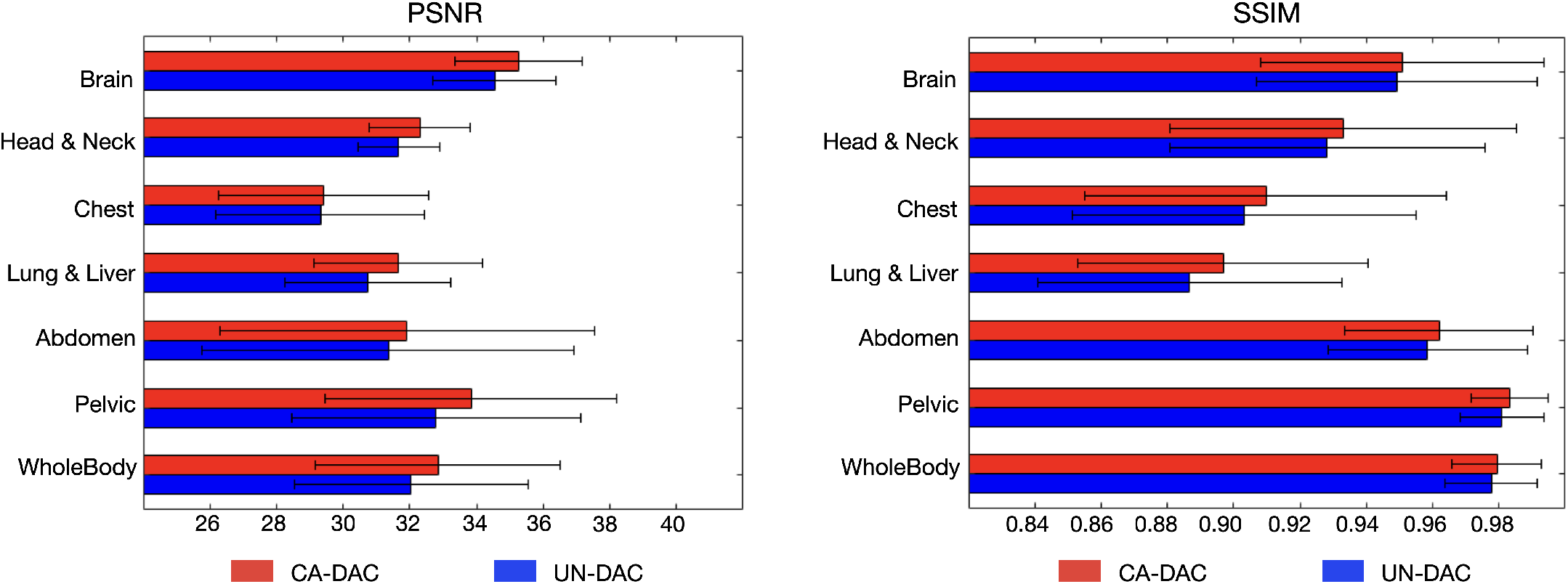
Quantitative error measured in 6 anatomical regions and whole-body for CA-DAC and UN-DAC approaches over 112 heldout subjects in terms of left) PSNR and right) SSIM.

We performed a joint histogram analysis to examine the correlation between the distribution of the corrected SUV values from the predicted PET-AC images and that of the reference PET-CT ground truth. The analysis was carried out within an SUV range of 0.5 to 12 on a log scale. As shown in Fig. 5, CA-DAC yields higher accuracy for lower uptake voxels as the distribution of the correlations exhibits less variation around the identity line. However, the correction accuracy starts to drop for both CA-DAC and UN-DAC in the voxels with higher uptake. The coefficient of determination, denoted by *R*^2^ was further used to quantify the goodness of fit within the joint histograms. In particular, *R*^2^ for CA-DAC and UN-DAC was recorded as 0.982 and 0.963, respectively, indicating a better fit using the former, i.e. proposed method. Moreover, we performed a linear regression analysis by reporting the slope and intercept of the line fitted over the non-zero bins of the joint histograms. The best performance is achieved when the slope and intercept equal to 1.0 and 0.0, respectively. As shown in Fig .5, UN-DAC yielded 0.81 and 1.30 for the slope and intercepts, respectively, while CA-DAC results in 0.88 and 0.57 demonstrating an increase of 0.07 for the slope (closer to 1.0) and a decrease of 0.73 for the intercept (closer to 0.0).

**Fig. 5.**
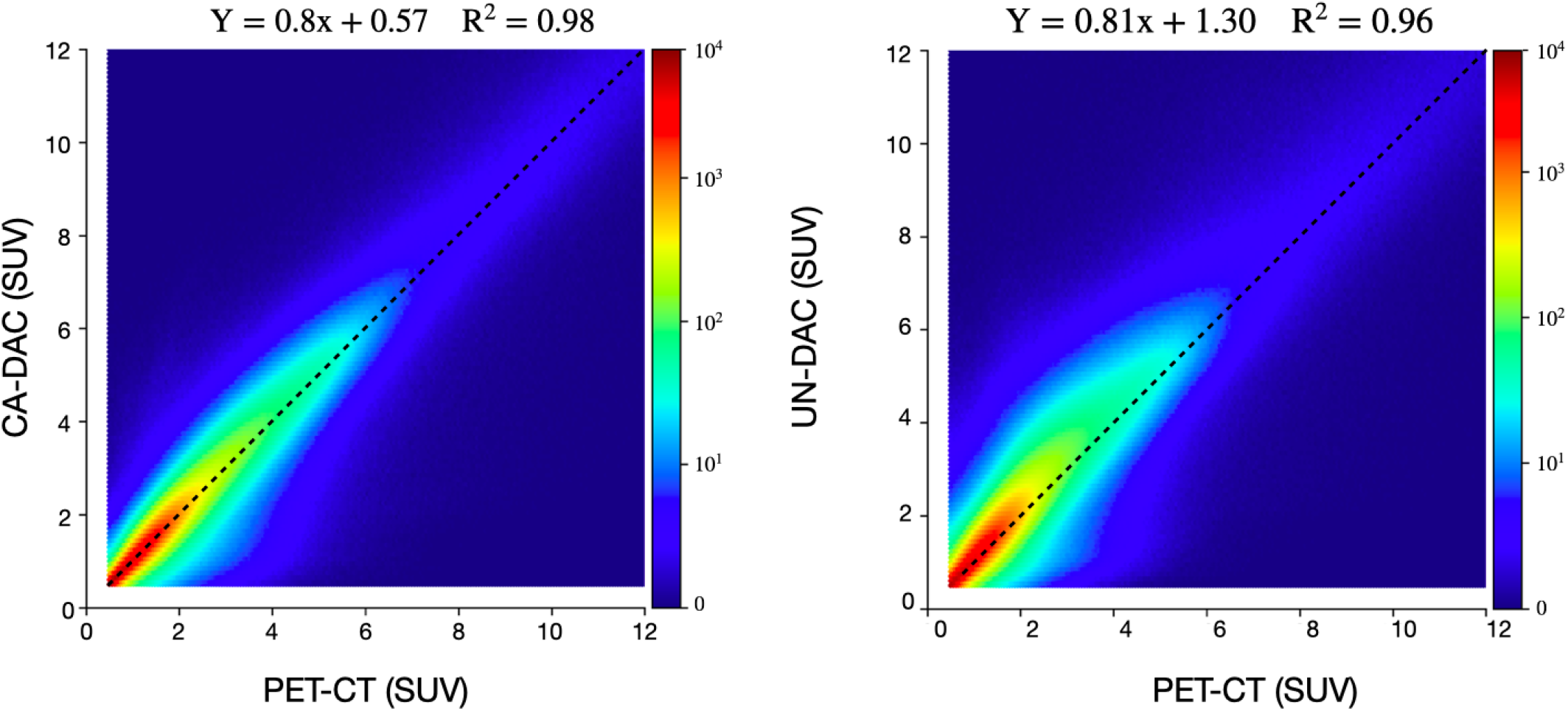
Joint histogram analysis displaying the correlation between PET-ASC images from left) CA-DAC and right) UN-DAC versus reference PET-CT ground truth. Note that a logarithmic scale was used to display the SUV levels

In the next analysis, we studied the impact of context-aware convolution in adapting the behavior of the network to the intra-subject uptake variations. Therefore, we approximately extracted 6 anatomical regions for every subject in the training set using empirical proportional presets over the whole-body image (brain: 10%, head & neck: 15%, chest: 20%, lung & liver: 5%, abdomen: 15%, and pelvic: 35%). We then trained 6 different 2.5D networks for each specific region over the entire training cohort (RGN-DAC) and evaluated them on a region-specific held-out cohort consisting of 50 subjects randomly selected from the primary 112 subjects. These networks provided us with an approximation of the upper-bound performance that could be achieved for each region. We then compared the performance of RGN-DAC against UN-DAC and CA-DAC models trained over whole-body images. As expected, RGN-DAC and UN-DAC yield the lowest and highest absolute relative errors, respectively, as shown in Table 2. However, we observed that context-aware convolutions in CA-DAC successfully enables the the network to outperform UN-DAC by 0.84 in terms of absolute relative error while still lagging behind RGN-DAC by 0.73 averaged over all regions.

**Table 2.** PET quantification error measure across 6 regions for UN-DAC, CA-DAC and RGN-DAC

| Region | UN-DAC | CA-DAC | RGN-DAC |
| --- | --- | --- | --- |
| Brain | $18.83 \pm 5.92$ | $17.12 \pm 4.82$ | $16.23 \pm 4.62$ |
| Head & Neck | $20.73 \pm 4.82$ | $20.04 \pm 4.22$ | $19.27 \pm 5.81$ |
| Chest | $23.23 \pm 6.61$ | $22.92 \pm 6.87$ | $22.42 \pm 6.34$ |
| Lung & Liver | $21.92 \pm 6.21$ | $21.34 \pm 5.69$ | $21.01 \pm 5.76$ |
| Abdomen | $18.48 \pm 4.01$ | $17.59 \pm 4.81$ | $16.36 \pm 4.32$ |
| Pelvic | $19.66 \pm 5.82$ | $18.83 \pm 6.11$ | $17.94 \pm 5.61$ |

Table 3 provides the image quality error computed on whole-body images in three different training schemes, namely UN-DAC (2D) that takes individual slices as input, UN-DAC (3D) which applies 3D convolutions on 3D volumetric patches of size (64 × 64 × 64), and our proposed CA-DAC, which takes individual 2D slices in the input but adapts the convolution kernels via contextual information. Among the different approaches, UN-DAC (3D) resulted in the largest number of parameters (19M) as it utilized 3D convolutions in the architecture. However, it yielded 0.9792 for SSIM and 32.86 for PSNR, which are not superior than the scores obtained by our proposed CA-DAC, i.e. 0.97936 for SSIM and 32.97 for PSNR. On the other hand, UN-DAC (2D) achieved the lowest scores for SSIM (0.9768), lowest PSNR (32.10), and the smallest number of parameters (7.1M). Compared to UN-DAC, we achieved superior accuracy with negligible increase in model complexity (200K more parameters).

**Table 3.** PET quantification error measured in whole-body for UN-DAC (2D), UN-DAC (3D), and CA-DAC in terms of SSIM and PSNR.

| Training Scheme | SSIM | PSNR | # Params |
| --- | --- | --- | --- |
| UN-DAC (2D) | $0.9768 \pm 0.01$ | $32.10 \pm 4.52$ | 7.1M |
| UN-DAC (3D) | $0.9791 \pm 0.01$ | $32.86 \pm 4.01$ | 19M |
| CA-DAC | $0.9793 \pm 0.01$ | $32.97 \pm 3.79$ | 7.3M |

### 4.2 Qualitative Evaluation

For qualitative inspection, we present a comparison of the visual results by UN-DAC and CA-DAC in Fig. 6. It was observed that both of these approaches produced visually indistinguishable results to the naked eyes (but not to SSIM and PSNR) compared to the reference PET-CT. However, as highlighted in section 4, a thorough comparison of their corresponding quantification error maps indicated that UN-DAC over-estimates SUV values in different regions, particularly in the abdomen region. Notably, AC methods usually produce considerable artifacts in the liver dome area due to the local mismatch between the PET and CT images caused by respiratory motion during the scan time. However, the error maps in Fig. 6 showed the our novel CAC layer succeeded to alleviate this problem in this error-prone region compared to UN-DAC. Lastly, Fig. 6 also presents the comparison of the horizontal profile drawn through the abdomen region for PET-CT and the PET-AC images estimated by CA-DAC and UN-DAC. The profile line by CA-DAC accurately matched the one by the reference PET-CT while the one by UN-DAC produced over-estimated SUV measurements for both low and high uptake areas. The qualitative results shown in Fig. 6 were consistent with the quantitative comparisons, which further demonstrates the effectiveness and efficiency of our proposed framework for attenuation correction.

**Fig. 6.**
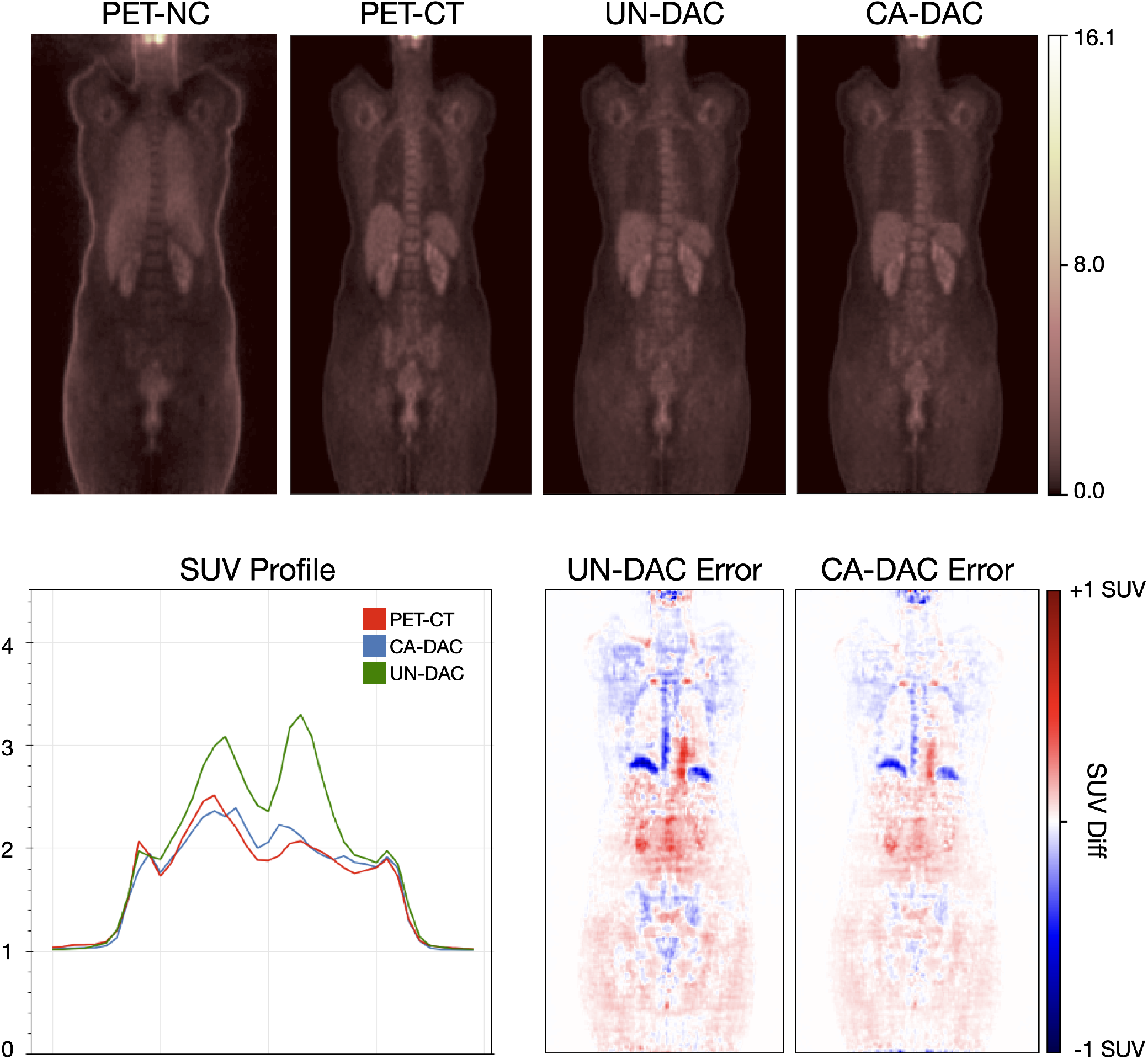
Qualitative comparison of coronal views of the PET images corrected for attenuation and scatter using UN-DAC and CA-DAC. Top panel provides the PET-NC image, PET-CT ground truth and PET-AC images obtained by UN-DAC and CA-DAC from left to right. Bottom right depicts the error map for UN-DAC and CA-DAC computed against PET-CT ground truths. Bottom left illustrates the tracer uptake profile along a line in abdomen region. A warm color map was chosen to improve the visualization of PET images.

## 5. Discussion

In clinical practice, automatic attenuation and scatter correction is an essential yet challenging problem in accurate PET quantification. CT and MRI related artifact can impose significant image quality degradation and/or artifacts on PET images. Artifacts present in anatomical imaging (CT and MRI) may propagate into corresponding PET-AC images. Respiratory mismatches in the thorax between PET and CT/MR images induce banana artifacts in PET images. Metallic and contrast agent material leads to strong streak artefacts and void signal in CT and MRI, respectively which change information of tissue attenuation/scatter factor, which results in inaccurate attenuation/scatter correction of PET images. Truncation artifacts of CT and MRI images appear in patients whose body size exceeds the transaxial field of view of MRI/CT images resulting in erroneous PET quantification. These errors, which propagate to the PET images in the ASC steps of the reconstruction, may not be easily identified due to the absence of a “ground truth”. Finally, use of compact PET-only scanners (e.g. for screening purposes) is a possibility in the future if attenuation and scatter correction issues, relying purely on PET images, can be addressed.

Many algorithms, including traditional and deep learning-based methods, have been explored to improve the PET quantification by correcting for attenuation/scatter artifacts. However, these methods failed to reach an efficient trade-off between exploiting the contextual information within neighboring slices and reducing the computational burden. Also, previous methods could not robustly adapt their performance in the presence of intra- and inter-subject uptake variations leading to inferior performance during inference. In this work, we proposed to learn a mapping between individual 2D slices in the PET-NC and PET-AC domains without access to anatomical information provided by either CT or MR. We further suggested augmenting the network architecture with CAC layers which led to a considerable performance gain as evidenced by the quantitative and qualitative results. We further utilized a massive cohort consisting of whole-body images from 910 subjects for training and evaluation.

Some stochastic interaction that causes photons to deviate from a straight line contributes to the attenuation/scatter artifacts. In contrast, the tracer uptakes within adjacent slices are highly correlated and can manifest qualitative and quantitative complementary patterns to each other. Therefore, incorporating the contextual information within neighboring slices into the network can efficiently suppress the impact of random and scatter coincidences recorded during PET image acquisition. In particular, the attenuation and scatter maps inferred in CAC layers can determine how the intermediate representations need to be filtered to accurately restore the PET-AC counterparts. Multiple inputs alone may not suffice to achieve an accurate PET-AC output. Thus, our proposed CA-DAC provides a more flexible ASC algorithm that firstly restores the missing details in every input slice through the inferred dynamic filters and then delivers the final PET-AC estimate for reference slide through eliminating the left over artifact using an averaging operation along slice dimension.

Through extensive quantitative analysis of the obtained PET-AC images (Fig 3 and Fig 4), we showed that our proposed CA-DAC network provided more accurate and robust SUV quantification in different anatomical regions compared to the baseline. The qualitative assessment of the predicted PET-AC images obtained by our proposed CA-DAC (Fig 6) revealed that it could achieve the results with enhanced visual quality, particularly in preserving detailed textures and removing sophisticated noise solely from PET-NC inputs. More importantly, visual inspections of the error maps in Fig 6 showed that our proposed CA-DAC could slightly alleviate the quantification error caused by mismatch artifacts in the liver dome region, which is most vulnerable to diaphragm and heart motion. Such improvements are important as it ameliorates the treatment of patients who cannot hold their breath well or in patients who have lesions in the liver dome. In future studies, we will investigate the capability of CA-DAC to address other common artifacts in PET imaging including radio-tracer related (halo artifact), patient related (motion, mismatch,and metal) and instrument related (truncation) artifacts (Shiri et al., 2022). In particular, out attempt will be focused on designing a efficient and fast quality assessment tool with the major objective of detecting and providing quantitative information in regions affected by common artifacts in PET imaging.

## 6. Conclusion

In this work, we introduced CA-DAC to produce attenuation-corrected PET images without requiring any anatomical information during training and inference. The novelty of the proposed network is to take advantage of context-aware convolutions to modulate the convolution kernels based on the contextual information within neighboring slices along the axial dimension for every 2D input slice. This way, the network can effectively adapt itself to the inter- and intra-subject tracer uptake variations with negligible increase in model complexity. The quantitative and qualitative results indicated considerable performance gain across the whole-body and 6 anatomical regions. This algorithm could be potentially used and evaluated in the context of different PET image artifacts detection and correction including patient and instrument related artifacts.

## Data Availability

All data produced in the present study are available upon reasonable request to the authors

## CRediT authorship contribution statement

Saeed Izadi: Conceptualization, Methodology, Writing - original draft. Isaac Shiri: Data curation, Writing - original draft. Carlos: Data curation. Parham Geramifar: Data curation, Habib Zaidi: Clinical supervision, Writing - original draft, Arman Rahmim: Clinical supervision, Writing - original draft. Ghassan Hamarneh: Technical supervision, Writing - original draft.

## 7. Acknowledgements

This work was supported by the Swiss National Science Foundation under Grant No. SNSF 320030 176052.

